# Patterns of peritoneal dissemination and response to systemic chemotherapy in common and rare peritoneal tumors treated by cytoreductive surgery: Study protocol of a prospective, multi-center, observational study

**DOI:** 10.1101/2021.04.01.21254760

**Authors:** Aditi Bhatt, Pascal Rousset, Dario Baratti, Daniele Biacchi, Nazim Benzerdjeb, Ignace de Hingh, Marcello Deraco, Vadim Gushcin, Praveen Kammar, Daniel Labow, Edward Levine, Brendan Moran, Faheez Mohamed, David Morris, Sanket Mehta, Aviram Nissan, Mohammad Alyami, Mohammad Adileh, Shoma Barat, Almog Ben Yacov, Kurtis Campbell, Kathleen Cummins-Perry, Delia Cortes-Guiral, Noah Cohen, Loma Parikh, Samer Alammari, Galal Bashanfer, Anwar Alshukami, Kaushal Kundalia, Gaurav Goswami, Vincent Van de Vlasakker, Michelle Sittig, Paolo Sammartino, Armando Sardi, Laurent Villeneuve, Kiran Turaga, Yutaka Yonemura, Olivier Glehen

## Abstract

**Introduction:** Despite optimal patient selection and surgical effort, recurrence is seen in over 70% of patients undergoing cytoreductive surgery(CRS) for peritoneal metastases (PM). Apart from the peritoneal cancer index(PCI), completeness of cytoreduction and tumor grade, there are other factors like disease distribution in the peritoneal cavity, pathological response to systemic chemotherapy(SC), lymph node metastases and morphology of PM which may have prognostic value. One reason for the underutilization of these factors is that they are known only after surgery. Identifying clinical predictors, specifically radiological predictors, could lead to better utilization of these factors in clinical decision making and the extent of peritoneal resection performed for different tumors. This study aims to study these factors, their impact on survival and identify clinical and radiological predictors.

**Methods and analysis:** There is no therapeutic intervention in the study. All patients with biopsy proven PM from colorectal, appendiceal, gastric and ovarian cancer and peritoneal mesothelioma undergoing CRS will be included. The demographic, clinical, radiological, surgical and pathological details will be collected according to a pre-specified format that includes details regarding distribution of disease, morphology of PM, regional node involvement and pathological response to SC. In addition to the absolute value of PCI, the structures bearing the largest tumor nodules and a description of the morphology in each region will be recorded. A correlation between the surgical, radiological and pathological findings will be performed and the impact of these potential prognostic factors on progression-free and overall survival determined. The practices pertaining to radiological and pathological reporting at different centers will be studied.

**Ethics and dissemination:** The study protocol has been approved by the Zydus Hospital ethics committee (27^th^ July, 2020) and Lyon-sud ethics committee (A15-128). It is registered with the clinical trials registry of India (CTRI/2020/09/027709).

The results will be published in peer-reviewed scientific journals.

**Strength and limitations:**

- A prospective correlation between the radiological, surgical and pathological findings in patients undergoing CRS will be performed which has not been done before.
- Being prospective in nature it will also enable us to evaluate the impact of the current treatment practices on the clinical end-points
- There is fixed protocol for radiological and pathological evaluation for which there are no specific guidelines
- The data collection format will capture all the relevant data but this may affect compliance.
- Despite the large sample size planned for each primary site, the heterogeneity of treatment protocols may be a limiting factor while evaluating the impact on survival.

## 1.0 Introduction

A wide variety of primary tumors give rise to peritoneal metastases (PM). Over the last couple of decades, the management of PM has undergone a radical change. Cytoreductive surgery (CRS) with, or without, some form of intraperitoneal chemotherapy has led to a major increase in survival, and cure in some patients. [1] Recurrence after CRS, however, is common and occurs in around 70% of the patients irrespective of the primary tumor site. [2, 3, 4] For all types of peritoneal malignancies the peritoneal cancer index (PCI) and the completeness of cytoreduction (CC-score) have emerged as the most important prognostic factors and are used for selecting patients for surgery. [5] The third major factor is disease biology, previously determined mainly by the histological subtype and/or grade and in more recent times using one or more molecular markers (for e.g. KRAS, BRAF and MSI for colorectal PM, BRCA mutations for ovarian cancer). [6] The main role of molecular markers, however, is in selecting patients for systemic therapies. In colorectal PM this is largely because there is limited evidence regarding the benefit of surgery in patients with tumors expressing poor prognostic markers. Apart from the grade of the tumor, pathological factors have been underutilized in selecting patients for surgery. The prognostic impact of the pathological response to systemic chemotherapy (SC) has been demonstrated for both colorectal PM and ovarian cancer. [7, 8] This, however, has not been utilized in clinical decision making. One reason could be that such factors are known only after the surgery has been completed. There are gaps in our knowledge of patterns of disease distribution and morphology of PM and results of experimental studies have not been evaluated in the clinical setting. [9, 10] In our preliminary study, we found a high incidence of regional lymph node involvement related to both the primary tumor and peritoneal disease. [11] All these factors, determined on pathological evaluation, could have a prognostic impact and could influence both patient selection and the extent of surgery performed.

Similarly, imaging has been used for determining the disease extent pre-operatively. As expected there is under-prediction of the PCI due to limitations of modern imaging modalities like MRI in detecting tiny peritoneal nodules (<5mm). The other information that can be derived from imaging, such as morphology of PM and disease distribution in the peritoneal cavity, has not been adequately explored for its potential clinical implications. Surgical decision making regarding the extent of resection is made on intra-operative visualization of peritoneal disease which has been reported to be inaccurate in a large proportion of the patients. [12, 13] There is no consensus on the extent of peritoneal resection that should be performed for each primary tumor. [14] And early recurrence could be due to the failure to address all the occult disease effectively with surgery. A better understanding of the disease distribution and mechanisms of peritoneal dissemination may help in standardizing the extent of resection for different primaries.

Correlation of the clinical radiological, surgical and pathological findings could provide new insights about peritoneal disease distribution, mechanisms of spread and potential impact of these prognostic factors on the treatment of patients with PM.

Hence, this study has been conceived and is being conducted with the following goals

- To study patterns of peritoneal disease distribution, lymph node metastases, morphology of PM and pathological response to SC in patients undergoing CRS.
- To study the impact of these factors on survival.
- To identify clinical and radiological predictors of these factors by performing a correlation between the radiological, surgical and pathological findings.
- To study the existing practices related to evaluation of disease extent on imaging, intra-operatively and on pathology and the extent of peritoneal resection performed for each tumor.
- To study the practices related to pathological evaluation of CRS specimens.

## 2.0 Methods

This is a prospective, multi-centre observational study. There is no therapeutic intervention. All the patients with biopsy proven PM from colorectal, appendiceal, gastric and ovarian cancer or with peritoneal mesothelioma undergoing CRS with, or without, intraperitoneal chemotherapy will be included. Informed consent will be taken from all patients. The demographic, clinical, radiological, surgical and pathological details will be collected according to a pre-specified format that includes details regarding the distribution of disease, morphology of PM, regional node involvement and pathological response to SC.

There are currently no reporting guidelines for both imaging and pathological evaluation of CRS specimens. The information captured can vary from center to center though the parameters analyzed are the same. The reporting format in this study **(supplement 1)** includes calculation of the radiological PCI and other details like the sites and structures bearing the largest tumor nodules and a description of the morphology in each region. Similarly, the surgical findings will be documented in a systematic pre-specified manner. To ensure uniformity in the morphological description, a list of morphological features has been made for each for the radiological, surgical and pathological evaluations. This description has to be provided with the lesion score for each region of the PCI. The first 6 months of the study comprise a test phase in which teams will see the feasibility and compliance of this form of data capturing. Based on the inputs from the participating centers, if required, some modifications will be made in the format of data collection. During the second phase, there will be an addition to the protocol for pathological evaluation in which pathologists will be required to take additional sections from the ‘normal appearing’ peritoneum adjacent to tumor nodules. Centers in which it is not possible to follow this protocol will continue to follow the protocol of the first phase. The study flow-chart is in **figure 1**.

**Figure 1.**
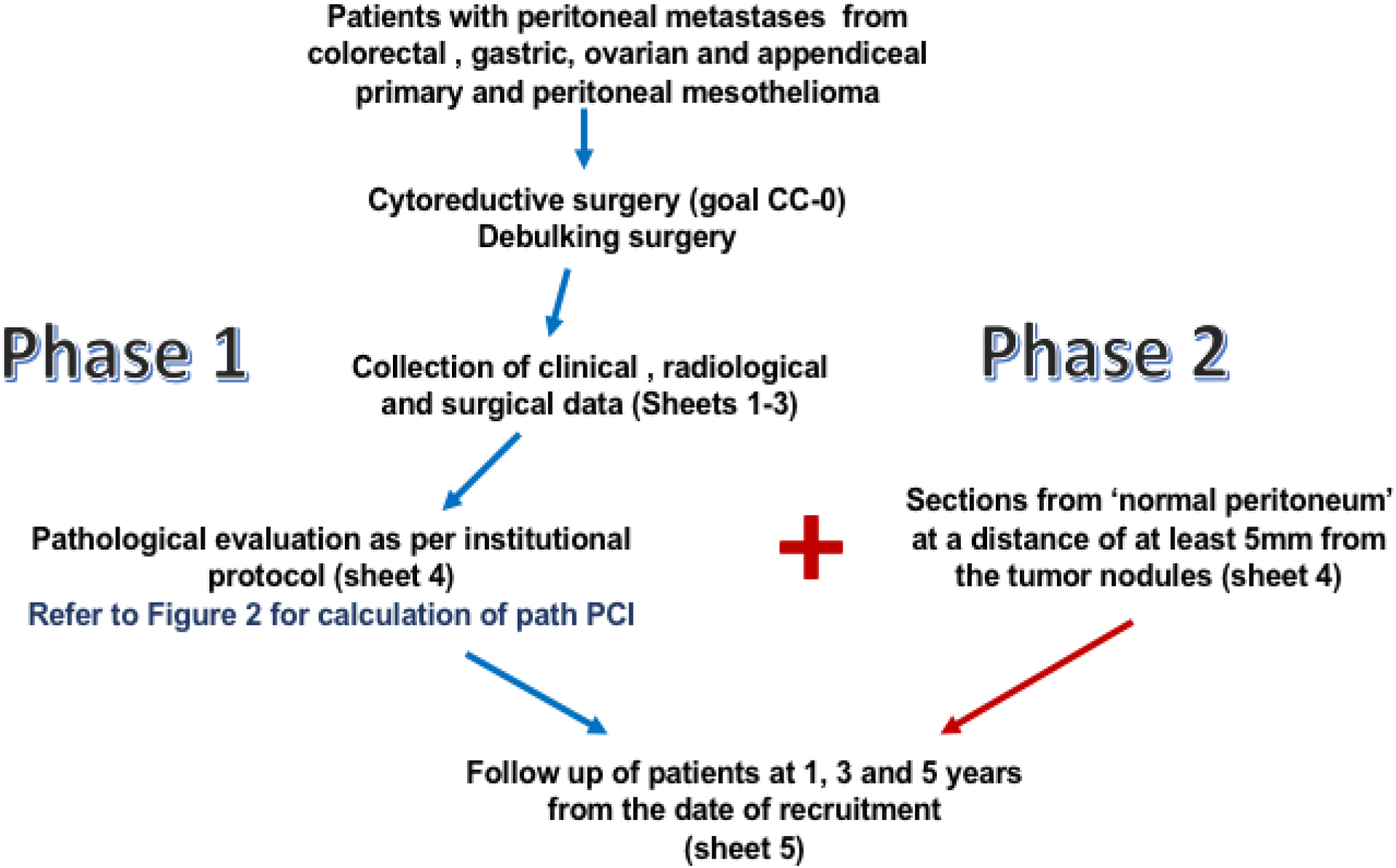
Study flow-chart and guide to data capturing

### 2.1 Inclusion and exclusion criteria

- All patients undergoing CRS for PM from colorectal, ovarian, gastric and appendiceal primary tumors or peritoneal mesothelioma will be included.
- Only patients with biopsy proven PM will be included (pathological evaluation is not mandatory prior to CRS if imaging or exploration confirms the presence of disease).
- Patients undergoing second look procedures with no evidence of peritoneal metastases will be excluded.
- Patient undergoing debulking procedures will be included
- Patients undergoing palliative procedures that do not involve tumor debulking will be excluded unless the goal of surgery was to obtain a complete cytoreduction i.e. procedures performed with the intention of palliation alone like a diverting stoma will be excluded.
- For ovarian cancer, only patients with FIGO stage III-C or IV-A will be included. Patients undergoing surgery at first diagnosis, after neoadjuvant chemotherapy (NACT) and those undergoing surgery for recurrent disease will be included. Patients without peritoneal disease will be excluded.

### 2.2 End points

#### Primary end points

The primary end point is disease distribution in the peritoneal cavity. Disease distribution in the peritoneal cavity will be captured in detail on imaging, during surgery and on pathology. The confirmation of disease on pathology will be considered as confirmatory for presence of disease in each region. If only debulking is performed, for regions which are not addressed during surgery, the surgical evaluation will be considered. The abdominal cavity is divided into 13 regions according to Sugarbaker’s PCI and the structures in each region are defined using the PROMISE internet application to ensure uniformity in reporting. [15, 16] Apart from the main data collection spreadsheet, separate forms have been created for the radiological, surgical and pathological evaluation to facilitation capturing of the disease distribution **(supplements 2-4)**.

#### Secondary end points

The secondary end-point are

1. **Pathological response to systemic chemotherapy**: This will be evaluated for all patients receiving NACT for PM. Different scores will be used for different primary tumors as described in the section on pathological evaluation.
2. **Regional lymph node involvement**: Both regional nodes in relation to the primary and those in relation to the PM are considered as regional nodes **(Supplement 1). [11]**
3. **Morphology of peritoneal metastases**: The morphological description for each region on imaging, during surgery and pathology will be documented **(Table 1)**. In phase two, presence of disease in adjacent normal peritoneum will be considered an additional morphological feature.
4. **Overall survival**: overall survival (OS) will be calculated from the date of surgery
5. **Progression-free survival**-progression-free survival (PFS) will be calculated from the date of surgery

**Table 1.** Morphological description of peritoneal lesions on radiological, intraoperative and pathological examination*

| Radiological evaluation | Visual inspection | Pathological examination |
| --- | --- | --- |
| Normal peritoneum | Normal peritoneum | Normal peritonuem |
| Tumor nodule/deposit | Tumor nodule | Tumor nodule |
| Scalloping | Confluent nodules | Clustering of nodules |
| Calcification | Plaque | Thickening |
| Thickening | Thickening | Scarring |
| Confluent disease | Scarring |  |
| Infiltration of adipose tissue | Adhesion |  |
| Retraction |  |  |

\*The description/definition of each morphological type is provided at the end of the PCI forms (Supplements 2-4)

### 2.3 Recruitment period

- 1^st^ Phase: 15^th^ Sept 2020-14th^st^ March 2021
- 2^nd^ Phase: 15^tt^ March 2021-14th^th^ Sept 2022

### 2.4 Study duration

The total duration of the study is 7 years; the first two years for recruitment and another 5 years for follow-up.

### 2.5 Surgical procedures

All surgical procedures will be performed with the goal of complete cytoreduction. [17] Patients undergoing planned debulking procedures will also be included. According to current surgical standards, only involved regions of the peritoneum or those bearing visible disease will be resected. Some regions like the falciform ligament, umbilical round ligament, lesser and greater omenta may be resected in the absence of visible disease as these structures have a high probability of harboring occult disease. These regions will be considered ‘normal appearing’ regions during surgical and pathological evaluation. At some centers, based on institutional policies and/or as part of an ongoing study, removal of the entire parietal peritoneum is performed for primary tumors such as mesothelioma, appendiceal and ovarian cancer to address occult disease. [18, 19] The peritoneal regions resected thus will be marked as ‘normal appearing’ regions.

The peritoneal lesions will be classified as nodules, plaques, confluent deposits, thickening, adhesions or scarring by the surgeon and given a lesion score accordingly **(Table 1)**. The largest deposit in each region of the PCI will have to be categorized thus, and the lesion score mentioned. The details of the peritonectomy procedures and visceral resections performed will be recorded.

### 2.6 Imaging protocol

CT scan with or without oral and iv contrast, FDG-PET CT scan (with IV contrast) and peritoneal MRI are all acceptable imaging modalities. A combination of the above may be performed. Where the facilities and expertise are available, peritoneal MRI is the preferred modality for evaluating the peritoneal disease. [20] Both T1 and T2, gadolinium enhanced and diffusion weighted images will be used to map the peritoneal disease. The protocol and sequences have been described in detail elsewhere and can be referred to. [20] When used in combination with MRI, oral contrast may be omitted while performing a CT scan. A slice thickness of 0.6-5mm is acceptable. If the whole thorax is not included, at least the lower thorax will be included.

Pre-operative imaging will be preferably performed within two to four weeks of the planning surgical procedures. All scans performed will be considered while evaluating both the presence or absence and extent of disease.

Imaging features suggestive of peritoneal metastases include peritoneal nodules, thickening, or fat stranding. The morphological classification is listed in Table 1.

There are two areas that are elaborated on here.

The first is disease related **peritoneal thickening**. Normal peritoneal tissues are relatively thin measuring <3 mm in thickness and typically show only no, or mild, enhancement that is less than or equal to that of the liver parenchyma. Obvious thickening, all the more if irregular or nodular, as well as marked peritoneal enhancement will be considered as peritoneal metastases (in proven cases of peritoneal metastases). Confluent nodules will be given a lesion score of 3 **(Supplement 2)**.

Infiltration of the adipose tissue (whatever the structure involved--mesentery –omentum – ligaments -mesocolon) will be be reported as “suggestive of peritoneal metastases” if at least confluent or pseudo-nodular in some areas, or nodular or mass-like. In the absence of diffuse mass-like disease scored 3 (e.g. omental cake), the lesion will be scored considering it as focal in the absence of measurable lesions. It will be scored 2 even if >5cm to avoid overestimation due to a misleading effect of resorption. If the maximum length of soft tissue is considered, it will be scored 2 if less than 5cm and 3 if more than 5cm.

### 2.7 Pathological evaluation

There are no guidelines or recommendations for evaluation of CRS specimens and each center will follow their existing protocols. [21] The evaluation should involve analysis of each peritoneal region submitted for evaluation separately. Similarly, all viscera should be evaluated individually. Centers where such an evaluation is not performed, or that cannot evaluate the specimens in this manner, were excluded from the study. One important aspect is calculation of the pathological PCI. [22] For the pathological PCI, the pathologist has to specify the size of the largest tumor deposit in the region in millimeters. The presence of disease in confirmed on microscopy. If there is no disease on microscopic examination, the lesion score is zero. The pathological response grade is mentioned for each of the thirteen regions. The method of calculating the pathological PCI is described in **Figure 2**.

**Figure 2.**
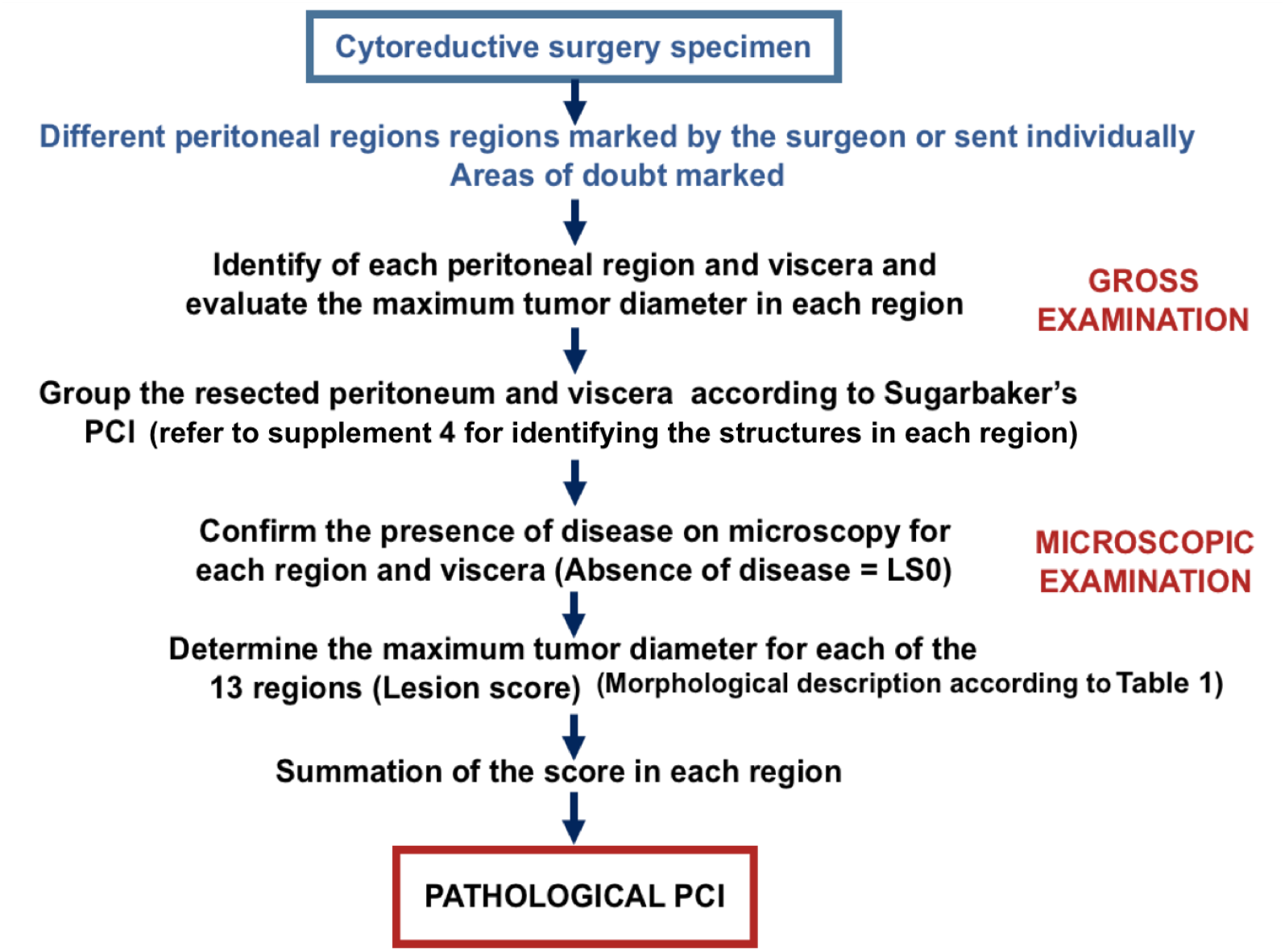
Calculation of the pathological PCI (from ref 22 with permission)

In the second phase, there will be an intervention in the pathological evaluation. For every region that is submitted to the pathologist, additional sections will be taken from the normal peritoneum at a distance of at least 5mm and preferably 10 mm from the tumor nodule. This evaluation is termed as evaluation of ‘normal peritoneum around tumor nodules’. This part of the study is not mandatory as institutional policies, time constraints and the cost involved may prevent centers performing this part of the study.

The following scores for evaluation for pathological response to SC will be used For ovarian cancer, the chemotherapy response score by Bohm et al has been validated in multiple studies and is the preferred score. It is described in **Table 2**. [23]

**Table 2.**
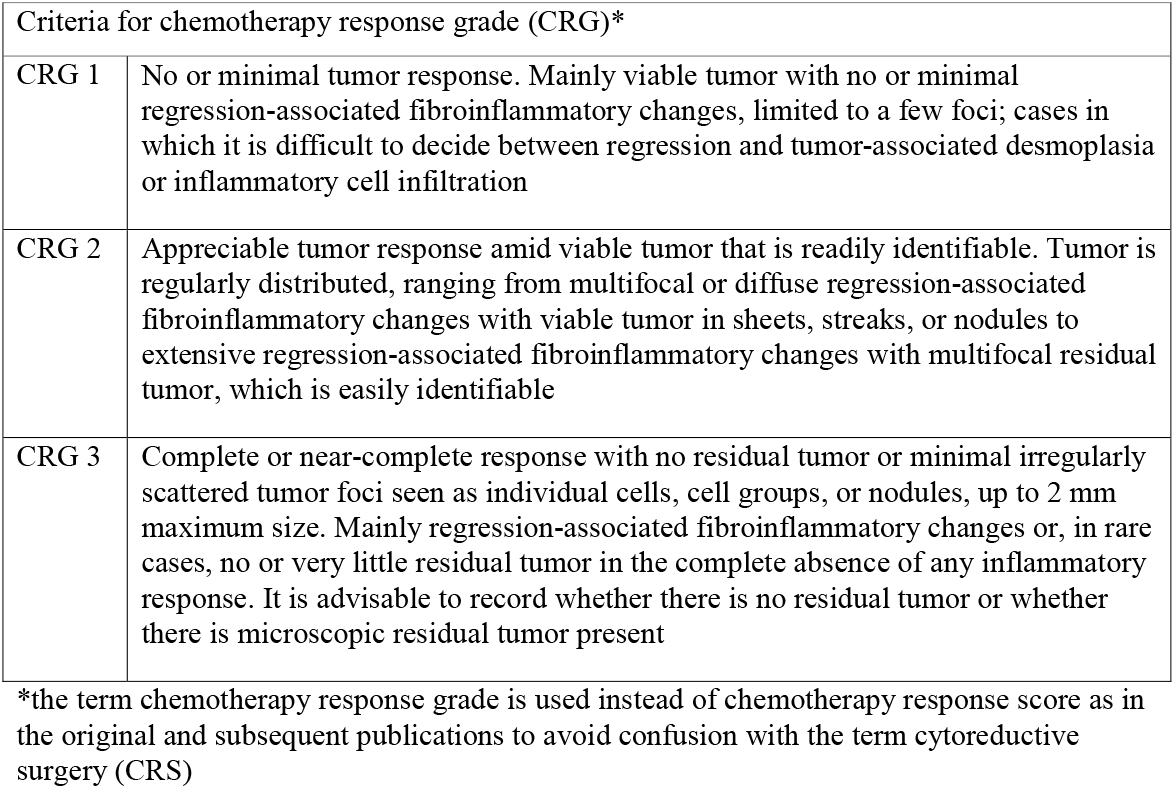
Score categorizing the pathological response to systemic chemotherapy in ovarian cancer [23]

| Criteria for chemotherapy response grade (CRG)* |  |
| --- | --- |
| CRG 1 | No or minimal tumor response. Mainly viable tumor with no or minimal regression-associated fibroinflammatory changes, limited to a few foci; cases in which it is difficult to decide between regression and tumor-associated desmoplasia or inflammatory cell infiltration |
| CRG 2 | Appreciable tumor response amid viable tumor that is readily identifiable. Tumor is regularly distributed, ranging from multifocal or diffuse regression-associated fibroinflammatory changes with viable tumor in sheets, streaks, or nodules to extensive regression-associated fibroinflammatory changes with multifocal residual tumor, which is easily identifiable |
| CRG 3 | Complete or near-complete response with no residual tumor or minimal irregularly scattered tumor foci seen as individual cells, cell groups, or nodules, up to 2 mm maximum size. Mainly regression-associated fibroinflammatory changes or, in rare cases, no or very little residual tumor in the complete absence of any inflammatory response. It is advisable to record whether there is no residual tumor or whether there is microscopic residual tumor present |

\*the term chemotherapy response grade is used instead of chemotherapy response score as in the original and subsequent publications to avoid confusion with the term cytoreductive surgery (CRS)

The peritoneal regression grading score by Solass et al was developed for patients undergoing PIPAC and has been externally validated **(Table 3)**. [24, 25] It can be used for any of the primary tumors.

**Table 3.**
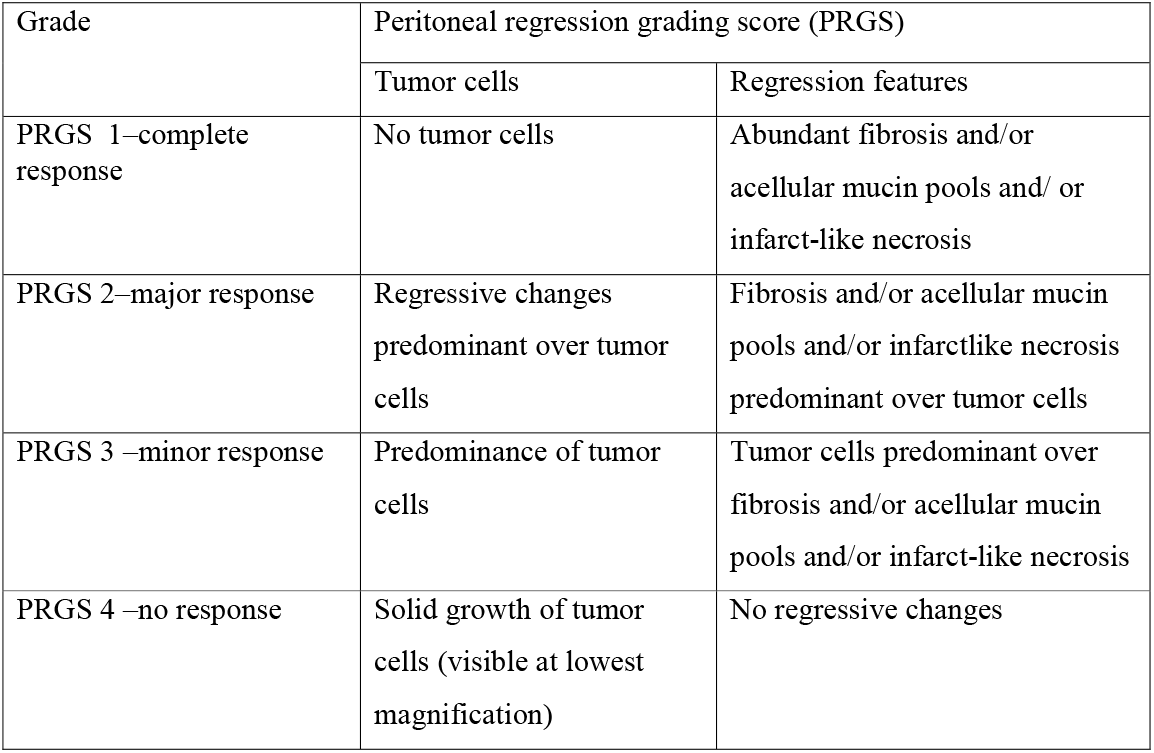
The peritoneal regression grading score (PRGS) that can be used for different primary tumours [24, 25]

| Grade | Peritoneal regression grading score (PRGS) |  |
| --- | --- | --- |
|  | Tumor cells | Regression features |
| PRGS 1–complete response | No tumor cells | Abundant fibrosis and/or acellular mucin pools and/ or infarct-like necrosis |
| PRGS 2–major response | Regressive changes predominant over tumor cells | Fibrosis and/or acellular mucin pools and/or infarctlike necrosis predominant over tumor cells |
| PRGS 3 –minor response | Predominance of tumor cells | Tumor cells predominant over fibrosis and/or acellular mucin pools and/or infarct-like necrosis |
| PRGS 4 –no response | Solid growth of tumor cells (visible at lowest magnification) | No regressive changes |

For colorectal PM, the score developed at Lyon-sud is an alternative as it is specific for colorectal PM but has not been externally validated **(Table 4)**. [26] The mean of scores in each region is computed and is the final score. In addition, the type of response is classified as fibrosis, infarct like necrosis or a colloid response. A combination of the three types of response can be present in a given region.

**Table 4.** The Lyon-sud score for colorectal PM [26]

| Grade of response |  | Type of response |
| --- | --- | --- |
| Complete response | No residual tumor cells | Fibrosis |
| Major response | 1-49% residual tumor cells | Infarct-like necrosis |
| Minor response | >50% residual tumor cells | Colloid response |

Use of scores apart from the above is permitted and the details of the same have to be provided. It would be ideal to have the same score for each primary at all centers but that again may not be possible. Secondly, the main focus will be a pathological complete response which does not have different criteria across the scores. Regional and peritoneal nodes will be evaluated as previously described.[11]

The reporting of other pathological findings like evaluation of the primary tumor, if present, grade or pathological classification will be according to the existing protocols at each center and will be captured in the data form. Similarly, the molecular markers that have been tested will be recorded with the test results. Performing specific molecular tests is not binding.

### 2.8 Follow up

The follow-up of patients will be performed according to the existing protocols at each centers. Centers will be asked to provide a follow-up of their patients at 1, 3 and 5 years after completion of the recruitment. The information will include the disease status, date of detection and site of recurrent disease, subsequent therapies administered, date and cause of death.

## 3.0 Sample size

The number of patients required was determined based on our preliminary study looking at the disease distribution in relation to the disease extent. We considered involvement of different regions in relation to the surgical PCI as well as different structures in relation to the surgical PCI for each primary tumor. In addition, consideration was given to another end-point, that is pathological response to SC. This was relevant only for colorectal cancer and ovarian cancer. For colorectal cancer, upper regions (1-3 of Sugarbaker’s PCI) were involved in 20%. [27] All patients with involvement of these regions had a surgical PCI of more than 10. [27] Thus, to confirm this finding in a larger series with 80% predictive power and alpha error of 0.05, we would require 35 patients with a PCI <10 and 35 with a PCI>10. Considering that 70% of the patients have a PCI <10 in those undergoing CRS, we would have to recruit over 100 patients. The incidence of involvement of the umbilical round ligament similarly, was 2% and we would have to recruit around 340 patients to confirm this. [27] For colorectal PM, few studies have evaluated the role of the pathological response to SC. The reported incidence of a pathological complete response (pCR) ranges from 10-15%. [26] In our recent analysis (unpublished data) it was 25%. In the same study, we could not find any factors that had a significant impact on the incidence of pCR except the type of chemotherapy used. To study the impact of 5-7 relevant factors we would need about 70 patients with a pCR. If we consider the incidence to be between 15 and 25, around 350 patients with colorectal PM receiving NACT will be required. Thus, the number of colorectal cancer patients we need to recruit in the study was set at 500 considering that around 70-80% will receive NACT.

For ovarian cancer, the incidence of a near complete/complete response or a Bohm score of 3 is around 15% in various published reports. [23] Many studies have explored predictors of a complete response and to date no factor has been found to have a significant impact. To determine the impact of 5 factors, 350 patients undergoing interval CRS alone would be needed. Considering the disease distribution, 120 patients undergoing primary CRS and CRS for recurrence each are required to be recruited. Thus 600 patients with ovarian cancer will be recruited. For peritoneal mesothelioma, 100 patients are required with numbers for gastric cancer being 100 patients and for appendiceal mucinous tumors/pseudomyxoma peritonei (PMP), 300 patients **(Table 5)**. These numbers also account for the variable histology, incidence of disease in different structures and regions that are crucial to studying the patterns of peritoneal dissemination.

**Table 5.** Sample size for each primary tumor site

|  |  |
| --- | --- |
| Colorectal cancer | 500 patients (350 patients who have received NACT) |
| Ovarian cancer | 600 patients (350 undergoing interval CRS) |
| Appendiceal primary | 300 patients |
| Peritoneal mesothelioma | 100 patients |
| Gastric cancer | 100 patients |

The study will continue for the stipulated time period even if the target numbers are recruited in a shorter period.

## 4.0 Statistical methods

Categorical data will be described as number (%). Non-normally distributed continuous data will be expressed as the median and range. Categorical data will be compared with the x2 test. For comparison of parametric data, the student t test and for non-parametric data, the Mann Whitney U test will be used. The impact of various prognostic variables like age, sex, primary tumour site, grade, primary tumour stage, PCI, timing of PM and NACT on the primary and secondary clinical end-points, namely disease distribution, pathological response to SC, morphology of PM and regional lymph node involvement will be studied. Multivariate logistic regression analysis will be performed to test the impact of multiple factors on these end-points. For studying the disease distribution, the peritoneal regions will be divided into four groups-upper regions comprising of regions 1,2,3, middle regions comprising of regions 0,4 and 8, lower regions comprising of regions 5, 6, 7 and small bowel regions comprising of regions 9-12 of Sugarbaker’s PCI. [16]

Previous studies have shown the negative prognostic impact of involvement of some regions like the upper regions and small bowel regions on survival.[28,29] The impact of involvement of these regions on survival will be evaluated as well. In addition, the involvement of some specific structures and regions like the omentum, right upper quadrant or region 1 will be considered separately.

Cox proportional hazard regression will be used to describe the association between individual risk factors, including the 4 pathological prognostic variables, on PFS and OS both, in terms of hazard ratio and its 95% confidence interval (CI). Multivariate Cox regression will be used to assess the impact of risk factors on survival. A p-value of <0.05 will be considered statistically significant. PFS and OS will be calculated from the date of surgery.

For identifying clinical predictors of these pathological prognostic factors, clinical and radiological variables will be considered. One of the important variables is the PCI (radiological and surgical). To test the performance of the radiological and surgical PCI, a comparison will be made with the pathological PCI for each of the two and also between the surgical and radiological PCI. A difference of 0-3 points will be considered as concordance. The comparison will be made between the total value and score in each region. The performance will be expressed in terms of false positives and negatives, true positives and negatives, sensitivity and specificity. Receiver operating characteristic (ROC) curves will be used to determine cut-off values of PCI to predict end-points like a pathological complete response.

PCI will also be evaluated as a categorical variable.

## 5.0 Ethics and dissemination

This study was approved by the Zydus hospital (institution of the first author) ethics committee on the 27^th^ July 2020. At Lyon-sud hospital, this study is being carried out within the framework of the RENAPE observational registry (CNIL-no. DR-2010-297) and the BIG-RENAPE registry (NCT NCT02823860), IRB number A15-128. [30] Subsequently, approval was obtained at other centres according to the existing institutional policies.

The proposed analyses will be carried out during the course and after completion of the study and the results published in peer-reviewed scientific journals.

## 6.0 Patient and public involvement

No patient involved

## 7.0 Registration

The study is registered with the clinical trials registry of India (CTRI/2020/09/027709).

## 8.0 Discussion

This study includes a prospective correlation between the radiological, surgical and pathological findings in patients undergoing CRS. The main goal is to determine prognostic impact of factors like disease distribution in the peritoneal cavity, pathological response to SC, lymph node metastases and morphology of PM. Correlation with clinical and radiological evaluation will be performed to identify predictors. The results could have a bearing, not just on the patient selection for surgery, but also the extent of peritoneal resection performed. Currently, the same surgical principles are applied to PM arising from different primary tumors, largely based on the surgical principles that were initially developed for pseudomyxoma peritonei (PMP) for which these procedures were first performed [31] It has been proposed that the surgical strategy should differ according to the primary tumor site. [32] The prognostic impact of pathological response to SC, involvement of specific regions of the peritoneal cavity and regional node involvement has been demonstrated previously for some primaries. [23, 33, 34] This study will look at these factors prospectively for all primary sites. This will be the first study to correlate the morphology on imaging and visual inspection with presence, or absence, of disease on pathology. For example, previous studies have looked at the false negative and false positives comparing the surgical lesion score and pathological finding. [35, 36] However, different surgeons will score different morphological appearances differently; some may give a lesion score of 0 to scarring, others may score it 1. The pathological PCI is a potential prognostic marker and is not calculated by many centers. If its prognostic value is demonstrated in this study, a simplified format of calculating it will be needed for other centers to be able to compute it. The results will provide information currently missing in the scientific literature regarding disease distribution in the peritoneal cavity in different primary tumors. It will differ from previous reports as a correlation with the PCI will be performed. Similarly, there is little information available about the regional lymph node involvement in relation to peritoneal disease. Nodal involvement in patients undergoing CRS can be in relation to the primary tumor if the primary is in situ or secondary to peritoneal disease. [11] Pelvic nodes, hepatic hilar nodes, greater and lesser omental nodes are some regional nodes that are involved secondary to peritoneal disease. Whilst regional nodes form part of staging of peritoneal mesothelioma, their involvement has not been studied in other tumors though surgeons often resect these nodes during CRS. Based on the results, the extent of peritoneal resection and regional lymphadenectomy that is performed can be determined for each primary tumor. The participating centers are some of the most experienced centers in treating peritoneal metastases and thus, though different treatment strategies will be employed at each center, their impact can be studied.

The format for data collection is exhaustive and designed to capture all relevant information. This may, however, become a limitation of the study as compliance will be a problem. The initial protocol underwent several modifications based on the inputs from each center before being finalized. Another review will be performed during the first phase and alterations made in the format of capturing data if the need arises. Despite the large sample size planned for each primary site, the heterogeneity of treatment protocols may be a limiting factor while evaluating the impact on survival. Nevertheless, the results will provide important insights on disease biology that will in the future influence the way these patients are treated.

Future studies could be performed on more homogeneous cohorts to validate the prognostic factors identified in this study.

## Data Availability

Study protocol- not applicable

## Author contributions

### Study design and conceptualization

Aditi Bhatt, Olivier Glehen

### Protocol writing

Aditi Bhatt, Olivier Glehen, Pascal Rousset, Nazim Benzerdjeb

### Data acquisition and analysis

Aditi Bhatt, Pascal Rousset, Dario Baratti, Daniele Biacchi, Nazim Benzerdjeb, Ignace de Hingh, Marcello Deraco, Vadim Gushcin, Praveen Kammar, Daniel Labow, Edward Levine, Brendan Moran, Faheez Mohamed, David Morris, Sanket Mehta, Aviram Nissan, Mohammad Alyami, Mohammad Adileh, Shoma Barat, Almog Ben Yacov, Kurtis Campbell, Kathleen Cummins-Perry, Delia Cortes-Guiral, Noah Cohen, Loma Parikh, Samer Alammari, Galal Bashanfer, Anwar Alshukami, Kaushal Kundalia, Gaurav Goswami,Vincent Van de Vlasakker, Michelle Sittig, Paolo Sammartino, Armando Sardi, Laurent Villeneuve, Kiran Turaga, Yutaka Yonemura, Olivier Glehen

### Interpretation of data

Aditi Bhatt, Pascal Rousset, Dario Baratti, Daniele Biacchi, Nazim Benzerdjeb, Ignace de Hingh, Marcello Deraco, Vadim Gushcin, Praveen Kammar, Daniel Labow, Edward Levine, Brendan Moran, Faheez Mohamed, David Morris, Sanket Mehta, Aviram Nissan, Mohammad Alyami, Mohammad Adileh, Shoma Barat, Almog Ben Yacov, Kurtis Campbell, Kathleen Cummins-Perry, Delia Cortes-Guiral, Noah Cohen, Loma Parikh, Samer Alammari, Galal Bashanfer, Anwar Alshukami, Kaushal Kundalia, Gaurav Goswami,Vincent Van de Vlasakker, Michelle Sittig, Paolo Sammartino, Armando Sardi, Laurent Villeneuve, Kiran Turaga, Yutaka Yonemura, Olivier Glehen

### Statistical calculations

Praveen Kammar, Aditi Bhatt and Olivier Glehen

### Critical review of the protocol and modifications

Dario Baratti, Marcello Deraco, Ignace de Hingh, Daniel Labow, Edward Levine, David Morris, Brendan Moran, Faheez Mohamed, Sanket Mehta, Aviram Nissan, Armando Sardi, Paolo Sammartino, Kiran Turaga, Yutaka Yonemura

### Ethics and permissions

Aditi Bhatt, Laurent Villeneuve, Faheez Mohamed, Michelle Sittig, Kathleen Cummins Perry, Shoma Barat, Mohammad Alyami

### Manuscript writing

Aditi Bhatt, Olivier Glehen

### Manuscript review and editing

Aditi Bhatt, Pascal Rousset, Dario Baratti, Daniele Biacchi, Nazim Benzerdjeb, Ignace de Hingh, Marcello Deraco, Vadim Gushcin, Praveen Kammar, Daniel Labow, Edward Levine, Brendan Moran, Faheez Mohamed, David Morris, Sanket Mehta, Aviram Nissan, Mohammad Alyami, Mohammad Adileh, Shoma Barat, Almog Ben Yacov, Kurtis Campbell, Kathleen Cummins-Perry, Delia Cortes-Guiral, Noah Cohen, Loma Parikh, Samer Alammari, Galal Bashanfer, Anwar Alshukami, Kaushal Kundalia, Gaurav Goswami,Vincent Van de Vlasakker, Michelle Sittig, Paolo Sammartino, Armando Sardi, Laurent Villeneuve, Kiran Turaga, Yutaka Yonemura, Olivier Glehen

### Approval of the final version of the manuscript

Aditi Bhatt, Pascal Rousset, Dario Baratti, Daniele Biacchi, Nazim Benzerdjeb, Ignace de Hingh, Marcello Deraco, Vadim Gushcin, Praveen Kammar, Daniel Labow, Edward Levine, Brendan Moran, Faheez Mohamed, David Morris, Sanket Mehta, Aviram Nissan, Mohammad Alyami, Mohammad Adileh, Shoma Barat, Almog Ben Yacov, Kurtis Campbell, Kathleen Cummins-Perry, Delia Cortes-Guiral, Noah Cohen, Loma Parikh, Samer Alammari, Galal Bashanfer, Anwar Alshukami, Kaushal Kundalia, Gaurav Goswami,Vincent Van de Vlasakker, Michelle Sittig, Paolo Sammartino, Armando Sardi, Laurent Villeneuve, Kiran Turaga, Yutaka Yonemura, Olivier Glehen We affirm that all the authors have provided substantial contribution to this manuscript and are in agreement with all aspects of the final version. All the authors agree to be accountable for all aspects of the work in ensuring that questions related to the accuracy or integrity of any part of the work are appropriately investigated and resolved.

