## Supplement 1 for "Patterns of peritoneal dissemination and response to systemic chemotherapy in common and rare peritoneal tumors treated by cytoreductive surgery: Study protocol of a prospective, multi-center, observational study"

Data collection form

Section 1: Demography, diagnosis, neoadjuvant treatment

| |
|---|
| Demographic data and previous therapies |
| Name/identifier |
| Centre |
| Country |
| Age |
| Sex |
| Primary tumor site |
| For colorectal cancer-exact site of the primary |
| For ovarian cancer- exact site (ovaries, FT, peritoneum) |
| Timing of PM (synchronous/metachronous) |
| Date of diagnosis of the primary |
| Date of diagnosis of PM |
| TNM stage at diagnosis |
| FIGO stage at diagnosis (for ovarian cancer) |
| Prior CRS |
| Date of prior CRS |
| Prior CRS+HIPEC |
| Date of prior CRS+HIPEC |
| Prior surgery (not CRS) |
| Prior surgical score |
| Previous chemotherapy |
| Number of lines of previous chemotherapy |
| Histological and molecular features |
| Histological diagnosis |
| Tumor grade (high/low) |
| Tumor differentiation (well, moderate, poor, undifferentiated) |
| Immunohistochemistry, if relevant (positive markers) |
| Immunohistochemistry, if relevant (negative markers) |
| BRCA (mutated/wt/not evaluated) |
| KRAS (mutated/wt/not evaluated) |
| BRAF (mutated/wt/not evaluated) |
| NRAS (mutated/wt/not evaluated) |
| BAP-1 (mutated/wt/not evaluated) |
| EGFR (mutated/wt/not evaluated) |
| Ki-67 (mention exact value in percentage) |

| |
|---|
| Any other marker (name) |
| Mutation status |
| Any other marker (name) |
| Mutation status |
| Neoadjuvant therapies |
| Neoadjuvant chemotherapy |
| Intravenous/oral |
| Regimen |
| Number of cycles |
| Date of starting |
| Date of completion |
| Neoadjuvant PIPAC |
| Regimen |
| Number of applications |
| Date of start |
| Date of completion |
| Neoadjuvant port directed IP chemotherapy |
| Regimen |
| Number of cycles |
| Date of start |
| Date of completion |
| Neoadjuvant HIPEC |
| Regimen |
| Number of cycles |

Section 2 Radiology

| |
|---|
| Peritoneal MRI |
| CT scan with oral and IV contrast |
| CT scan with IV contrast |
| PET CT scan |
| Any other (please specify) |
| Radiological PCI |
| Region 0 |
| Morphological description |
| Region 1 |
| Morphological description |
| Region 2 |
| Morphological description |
| Region 3 |

| |
|---|
| Morphological description |
| Region 4 |
| Morphological description |
| Region 5 |
| Morphological description |
| Region 6 |
| Morphological description |
| Region 7 |
| Morphological description |
| Region 8 |
| Morphological description |
| Region 9 |
| Morphological description |
| Region 10 |
| Morphological description |
| Region 11 |
| Morphological description |
| Region 12 |
| Morphological description |
| Evaluation of response |
| RECIST criteria |
| Any other(please specify) |
| Progressive disease |
| Stable disease |
| Partial disease |
| Complete response |
| Nodal disease |
| Right pelvic nodes (disease/no disease) |
| Max diameter |
| Left pelvic nodes (D/ND) |
| Max diameter |
| Paraaortic nodes (D/ND) |
| Max diameter |
| Mesorectal nodes (D/ND) |
| Max diameter |
| Paracolic nodes (D/ND) |
| Max diameter |
| Small bowel nodes (D/ND) |
| Max diameter |
| CBD/periportal (D/ND) |
| Max diameter |
| Left gastric nodes (D/ND) |

| |
|---|
| Max diameter |
| Peripancreatic nodes (D/ND) |
| Max diameter |
| Supradiaphragmatic nodes (D/ND) |
| Max diameter |
| Paracardiac (D/ND) |
| Max diameter |
| Any other-specify (D/ND) |
| Max diameter |
| Peritoneal nodes (D/ND) |
| Max diameter |
| Ascites |
| Ascites |
| Quantity |
| Characteristic |
| Density (for mucinous ascites) |
| Primary tumor site |
| Primary tumor site (evaluated/not evaluated) |
| Disease- present or absent |
| Response in the primary |
| Regional nodes |
| Radiological T stage |
| Radiological N stage |
| Radiological M stage |
| Previous anastomosis (evaluated/not evaluated) |
| Disease- present/absent |
| Stoma site- evaluated/not evaluated |
| Disease- present/absent |
| PORT site- evaluated/not evaluated |
| Disease-present/absent |
| Previous surgical scar- evaluated/not evaluated |
| Disease-present/absent |

Section 3 Surgical and perioperative details

| |
|---|
| ASA |
| ECOG Status |
| Date of CRS |
| Resection of the primary |

| |
|---|
| HIPEC |
| Open/closed |
| Drug regimen |
| Duration |
| Surgical PCI |
| Region 0 |
| Morphological description |
| Region 1 |
| Morphological description |
| Region 2 |
| Morphological description |
| Region 3 |
| Morphological description |
| Region 4 |
| Morphological description |
| Region 5 |
| Morphological description |
| Region 6 |
| Morphological description |
| Region 7 |
| Morphological description |
| Region 8 |
| Morphological description |
| Region 9 |
| Morphological description |
| Region 10 |
| Morphological description |
| Region 11 |
| Morphological description |
| Region 12 |
| Morphological description |
| Perioperative outcomes |
| Duration of surgery |
| Blood loss |
| Post op ventilation (hours) |
| ICU stay |
| Hospital stay |
| Grade 1-2 complications |
| 90-day grade 3-4 morbidity |
| Haemorrhage |
| Specify |
| Hematological toxicity |

| |
|---|
| Specify |
| Respiratory complications |
| Specify |
| Sepsis |
| Specify |
| Neutropenia |
| Lowest count |
| Cardiac complications |
| Specify |
| GI complications |
| Specify |
| Bowel fistula |
| Specify |
| Bowel perforation |
| Specify site |
| Renal complication |
| Specify |
| Nephrological complications |
| Specify |
| Wound dehiscence |
| Site |
| Surgical site infection |
| Return to ICU |
| Duration of stay |
| Return to operating room |
| Procedure performed/indication |
| Radiological intervention |
| Specify |
| Mortality |
| Date of death |
| Cause of death |

Section 4 Pathology

| |
|---|
| No of specimens |
| No of blocks |
| Pathological PCI |
| Region 0 |
| Morphological description |
| Region 1 |
| Morphological description |
| Region 2 |
| Morphological description |

| |
|---|
| Region 3 |
| Morphological description |
| Region 4 |
| Morphological description |
| Region 5 |
| Morphological description |
| Region 6 |
| Morphological description |
| Region 7 |
| Morphological description |
| Region 8 |
| Morphological description |
| Region 9 |
| Morphological description |
| Region 10 |
| Morphological description |
| Region 11 |
| Morphological description |
| Region 12 |
| Morphological description |
| Nodal disease |
| Right pelvic nodes (dissected/not dissected) |
| No of nodes |
| No of positive nodes |
| Left pelvic nodes (D/ND) |
| No of nodes |
| No of positive nodes |
| Paraaortic nodes (D/ND) |
| No of nodes |
| No of positive nodes |
| Mesorectal nodes (D/ND) |
| No of nodes |
| No of positive nodes |
| Paracolic nodes (D/ND) |
| No of nodes |
| No of positive nodes |
| Small bowel nodes (D/ND) |
| No of nodes |
| No of positive nodes |
| CBD/periportal (D/ND) |
| No of nodes |
| No of positive nodes |
| Left gastric nodes (D/ND) |
| No of nodes |

| |
|---|
| No of positive nodes |
| Peripancreatic nodes (D/ND) |
| No of nodes |
| No of positive nodes |
| Supradiaphragmatic nodes (D/ND) |
| No of nodes |
| No of positive nodes |
| Paracardiac (D/ND) |
| No of nodes |
| No of positive nodes |
| Any other-specify (D/ND) |
| No of nodes |
| No of positive nodes |
| Peritoneal nodes (D/ND) |
| No of nodes |
| No of positive nodes |
| Greater omental nodes |
| No of nodes |
| No of positive nodes |
| Lesser omental nodes |
| No of nodes |
| No of positive nodes |
| Total no of nodes |
| Total no of positive nodes |
| Pathological response to systemic chemotherapy |
| Bohm score(for ovarian cancer) |
| Response grade |
| PRGS |
| Response grade |
| Lyon-score for colorectal PM |
| Overall score |
| Fibrosis (%) |
| Necrosis (%) |
| Colloid response (%) |
| Any other score used |
| Grade |
| Additional comments |
| Pathological complete response (y/n) |
| Sites of complete response (present/absent) |
| Evaluation of the primary tumour |
| Disease in the primary |
| Maximum tumour diameter |

| |
|---|
| T-stage |
| N-stage |
| No of dissected nodes |
| No of positive nodes |
| Extracapsular spread |
| Lymphovascular invasion |
| Perineural invasion |
| Immunohistochemistry-positive markers |
| Immunohistochemistry-negative markers |
| Target regions and normal peritoneum |
| Disease in normal peritoneum |
| Falciform ligament (resected/not resected) |
| Max tumour diameter on gross examination (mm) |
| Microscopic disease (present/absent) |
| Umbilical round ligament (resected/not resected) |
| Max tumour diameter on gross examination (mm) |
| Microscopic disease (present/absent) |
| Greater omentum (resected/not resected) |
| Max tumour diameter on gross examination (mm) |
| Microscopic disease (present/absent) |
| Lesser omentum (resected/not resected) |
| Max tumour diameter on gross examination (mm) |
| Microscopic disease (present/absent) |
| Pancreatic capsule (resected/not resected) |
| Max tumour diameter on gross examination (mm) |
| Microscopic disease (present/absent) |
| Viscera |
| Glisson's (resected/not resected) |
| Max tumour diameter on gross examination (mm) |
| Microscopic disease (present/absent) |
| Rectum/rectosigmoid colon (resected/not resected) |
| Max tumour diameter on gross examination (mm) |
| Microscopic disease (present/absent) |
| Right colon (resected/not resected) |
| Max tumour diameter on gross examination (mm) |
| Microscopic disease (present/absent) |
| Gall bladder (resected/not resected) |
| Max tumour diameter on gross examination (mm) |
| Microscopic disease (present/absent) |
| Spleen (resected/not resected) |
| Max tumour diameter on gross examination (mm) |
| Microscopic disease (present/absent) |

| |
|---|
| Stomach (resected/not resected) |
| Max tumour diameter on gross examination (mm) |
| Microscopic disease (present/absent) |
| Small bowel (resected/not resected) |
| Max tumour diameter on gross examination (mm) |
| Microscopic disease (present/absent) |
| Liver (resected/not resected) |
| Max tumour diameter on gross examination (mm) |
| Microscopic disease (present/absent) |
| Intraparenchymal metastases (resected/not resected) |
| Max tumour diameter on gross examination (mm) |
| Microscopic disease (present/absent) |
| Previous anastomotic site (resected/not resected) |
| Max tumour diameter on gross examination (mm) |
| Microscopic disease (present/absent) |
| Previous stoma site (resected/not resected) |
| Max tumour diameter on gross examination (mm) |
| Microscopic disease (present/absent) |
| PORT SITE (resected/not resected) |
| Max tumour diameter on gross examination (mm) |
| Microscopic disease (present/absent) |
| Previous surgical scar (resected/not resected) |
| Max tumour diameter on gross examination (mm) |
| Microscopic disease (present/absent) |

Section 5 Adjuvant therapy and follow up

| |
|---|
| Adjuvant therapy |
| Adjuvant therapy planned |
| Date of start of adjuvant therapy |
| Chemotherapy regimen |
| Number of cycles |
| Targetted therapy |
| Drug |
| Number of cycles |
| Date of last cycle |
| Maintenance therapy |
| Drug |
| Duration |
| Follow-up |
| Last follow-up |

| |
|---|
| Disease status |
| Date of diagnosis of recurrence |
| Site of recurrence |
| Site of metastases |
| Treatment for recurrence/metastatic disease |
| Date of death |
| Cause of death |
