## Supplement 2 for "Patterns of peritoneal dissemination and response to systemic chemotherapy in common and rare peritoneal tumors treated by cytoreductive surgery: Study protocol of a prospective, multi-center, observational study"

Radiological PCI
Select the structure with the largest tumor deposit in the region, select the type of lesion and the score (any one in each column)

| Region | Structure | Morphology (select any one) | Lesion score |
|---|---|---|---|
| 0 | Midline incision [_]<br>Anterior parietal peritoneum [_]<br>Greater omentum [_]<br>Transverse colon [_]<br>Gastrocolic ligament [_]<br>Transverse mesocolon [_] | Normal peritoneum [_]<br>Tumour nodule [_]<br>Scalloping [_]<br>Calcification [_]<br>Thickening [_]<br>Confluent disease [_]<br>Infiltration of fat tissue [_]<br>Omental cake [_]<br>Retraction [_] | 0 [_]<br>1 [_]<br>2 [_]<br>3 [_] |
| 1 | Right hepatic lobe surface [_]<br>Diaphragm [_]<br>Foramen of Winslow [_]<br>Gallbladder [_]<br>Hepatorenal recess [_] | Normal peritoneum [_]<br>Tumour nodule [_]<br>Scalloping [_]<br>Calcification [_]<br>Thickening [_]<br>Confluent disease [_]<br>Infiltration of fat tissue [_]<br>Retraction [_] | 0 [_]<br>1 [_]<br>2 [_]<br>3 [_] |
| 2 | Left hepatic lobe surface [_]<br>Lesser omentum [_]<br>Hepatic hilum [_]<br>Falciform ligament [_] | Normal peritoneum [_]<br>Tumour nodule [_]<br>Scalloping [_]<br>Calcification [_]<br>Thickening [_]<br>Confluent disease [_]<br>Infiltration of fat tissue [_]<br>Retraction [_] | 0 [_]<br>1 [_]<br>2 [_]<br>3 [_] |
| 3 | Surface of spleen [_]<br>Left diaphragmatic peritoneum [_]<br>Tail of pancreas and hilum of the spleen [_]<br>Anterior and posterior stomach surfaces [_] | Normal peritoneum [_]<br>Tumour nodule [_]<br>Scalloping [_]<br>Calcification [_]<br>Thickening [_]<br>Confluent disease [_]<br>Infiltration of fat tissue [_]<br>Retraction [_] | 0 [_]<br>1 [_]<br>2 [_]<br>3 [_] |
| 4 | Left colon [_]<br>Left paracolic gutter [_]<br>Left mesocolon [_] | Normal peritoneum [_]<br>Tumour nodule [_]<br>Scalloping [_]<br>Calcification [_]<br>Thickening [_]<br>Confluent disease [_] | 0 [_]<br>1 [_]<br>2 [_]<br>3 [_] |

| | | | |
|---|---|---|---|
| | | Infiltration of fat tissue [_]<br>Retraction [_] | |
| 5 | Left pelvic peritoneum [_]<br>Secondary root of the mesosigmoid [_]<br>Sigmoid colon [_] | Normal peritoneum [_]<br>Tumour nodule [_]<br>Scalloping [_]<br>Calcification [_]<br>Thickening [_]<br>Confluent disease [_]<br>Infiltration of fat tissue [_]<br>Retraction [_] | 0 [_]<br>1 [_]<br>2 [_]<br>3 [_] |
| 6 | Uterus [_]<br>Left Fallopian tube [_]<br>Right Fallopian tube [_]<br>Left ovary [_]<br>Right ovary [_]<br>Bladder [_]<br>Pouch of Douglas [_]<br>Rectosigmoid junction [_] | Normal peritoneum [_]<br>Tumour nodule [_]<br>Scalloping [_]<br>Calcification [_]<br>Thickening [_]<br>Confluent disease [_]<br>Infiltration of fat tissue [_]<br>Retraction [_] | 0 [_]<br>1 [_]<br>2 [_]<br>3 [_] |
| 7 | Right pelvic peritoneum [_]<br>Cecum [_]<br>Appendix [_]<br>Mesoappendix [_] | Normal peritoneum [_]<br>Tumour nodule [_]<br>Scalloping [_]<br>Calcification [_]<br>Thickening [_]<br>Confluent disease [_]<br>Infiltration of fat tissue [_]<br>Retraction [_] | 0 [_]<br>1 [_]<br>2 [_]<br>3 [_] |
| 8 | Right colon [_]<br>Right paracolic gutter [_]<br>Right mesocolon [_] | Normal peritoneum [_]<br>Tumour nodule [_]<br>Scalloping [_]<br>Calcification [_]<br>Thickening [_]<br>Confluent disease [_]<br>Infiltration of fat tissue [_]<br>Retraction [_] | 0 [_]<br>1 [_]<br>2 [_]<br>3 [_] |
| 9 | Proximal jejunum [_]<br>Proximal mesojejunum [_] | Normal peritoneum [_]<br>Tumour nodule [_]<br>Scalloping [_]<br>Calcification [_]<br>Thickening [_]<br>Confluent disease [_]<br>Infiltration of fat tissue [_]<br>Mesenteric retraction [_] | 0 [_]<br>1 [_]<br>2 [_]<br>3 [_] |
| 10 | Distal jejunum [_]<br>Distal mesojejunum [_] | Normal peritoneum [_]<br>Tumour nodule [_]<br>Scalloping [_]<br>Calcification [_]<br>Thickening [_]<br>Confluent disease [_]<br>Infiltration of fat tissue [_]<br>Mesenteric retraction [_] | 0 [_]<br>1 [_]<br>2 [_]<br>3 [_] |

| | | | |
|---|---|---|---|
| 11 | Proximal ileum [_]<br>Proximal mesoileum [_] | Normal peritoneum [_]<br>Tumour nodule [_]<br>Scalloping [_]<br>Calcification [_]<br>Thickening [_]<br>Confluent disease [_]<br>Infiltration of fat tissue [_]<br>Mesenteric retraction [_] | 0 [_]<br>1 [_]<br>2 [_]<br>3 [_] |
| 12 | Distal ileum [_]<br>Distal ileum [_] | Normal peritoneum [_]<br>Tumour nodule [_]<br>Scalloping [_]<br>Calcification [_]<br>Thickening [_]<br>Confluent disease [_]<br>Infiltration of fat tissue [_]<br>Mesenteric retraction [_] | 0 [_]<br>1 [_]<br>2 [_]<br>3 [_] |
| | Total | | |

Key points

- Each structure is considered only in one region. For e.g., a large omental cake may extend beyond region 0, but should be scored only in region 0
- When assigning the lesion score, the score of the structure bearing the largest tumor deposit should be considered and the morphology of that lesion scored

- The morphological features are defined as follows

1. **Normal peritoneum**- Absence of any of the changes described below.

2. **Tumor nodule** : Elevated mass with a round or irregular appearance

3. **Scalloping-** A series of indentations or erosion on the smooth margin of a visceral structure (can be one or more). This is most commonly seen in case of mucinous peritoneal deposits on the diaphragmatic peritoneum that indent the surface of the liver or the spleen

4. **Calcification** Presence of one or more areas of calcification

5. **Thickening:** Normal peritoneal tissues are relatively thin measuring <3 mm in thickness and typically show only no or mild enhancement that is less than or equal to that of the liver parenchyma. Obvious thickening, especially if it is irregular or nodular as well as marked peritoneal enhancement should be considered as peritoneal metastases in a known case of peritoneal metastases.

6. **Confluent disease**: More than one nodule in a region with the intervening peritoneum showing abnormalities like smaller nodules or thickening. Alternatively, when the edges of discrete nodules become confluent.

7. **Infiltration of fat tissue**: Infiltration of the adipose tissue (whatever the structure involved-- mesentery -omenta – ligaments -mesocolon) is reported as suggestive of peritoneal metastases if at least confluent or pseudo-nodular in some areas, or nodular or mass-like. In absence of diffuse mass-like disease scored 3 (ex: omental cake), the lesion should be scored considering it as focal in absence of measurable lesion and score 2 even if >5cm to avoid overestimation due to a misleading part of resorption, or measuring the maximum length of soft tissue portion < 5cm – score 2 or > 5cm -score 3.

8. **Mesenteric retraction**: Tumor induced fibrosis and foreshortening of the mesentery leading to clumping of the bowel loops.
