## Supplement 3 for "Patterns of peritoneal dissemination and response to systemic chemotherapy in common and rare peritoneal tumors treated by cytoreductive surgery: Study protocol of a prospective, multi-center, observational study"

Surgical PCI (Select the structure with the largest tumor deposit in the region)

| Region | Structure | Morphology (select any one) | Lesion score |
|---|---|---|---|
| 0 | Midline incision [_]<br>Anterior parietal peritoneum [_]<br>Greater omentum [_]<br>Omental cake [_]<br>Transverse colon [_]<br>Gastrocolic ligament [_]<br>Transverse mesocolon [_] | Tumour nodule [_]<br>Confluent nodules [_]<br>Plaque [_]<br>Thickening [_]<br>Scarring [_]<br>Adhesion [_]<br>Normal peritoneum [_] | 0 [_]<br>1 [_]<br>2 [_]<br>3 [_] |
| 1 | Right hepatic lobe surface [_]<br>Diaphragm [_]<br>Foramen of Winslow [_]<br>Gallbladder [_]<br>Hepatorenal recess [_] | Tumour nodule [_]<br>Confluent nodules [_]<br>Plaque [_]<br>Thickening [_]<br>Scarring [_]<br>Adhesion [_]<br>Normal peritoneum [_] | 0 [_]<br>1 [_]<br>2 [_]<br>3 [_] |
| 2 | Left hepatic lobe surface [_]<br>Lesser omentum [_]<br>Hepatic hilum [_]<br>Falciform ligament [_] | Tumour nodule [_]<br>Confluent nodules [_]<br>Plaque [_]<br>Thickening [_]<br>Scarring [_]<br>Adhesion [_]<br>Normal peritoneum [_] | 0 [_]<br>1 [_]<br>2 [_]<br>3 [_] |
| 3 | Surface of spleen [_]<br>Left diaphragmatic peritoneum [_]<br>Tail of pancreas and hilum of the spleen [_]<br>Anterior and posterior stomach surfaces [_] | Tumour nodule [_]<br>Confluent nodules [_]<br>Plaque [_]<br>Thickening [_]<br>Scarring [_]<br>Adhesion [_]<br>Normal peritoneum [_] | 0 [_]<br>1 [_]<br>2 [_]<br>3 [_] |
| 4 | Left colon [_]<br>Left paracolic gutter [_]<br>Left mesocolon [_] | Tumour nodule [_]<br>Confluent nodules [_]<br>Plaque [_]<br>Thickening [_]<br>Scarring [_]<br>Adhesion [_]<br>Normal peritoneum [_] | 0 [_]<br>1 [_]<br>2 [_]<br>3 [_] |
| 5 | Left pelvic peritoneum [_]<br>Secondary root of the mesosigmoid [_]<br>Sigmoid colon [_] | Tumour nodule [_]<br>Confluent nodules [_]<br>Plaque [_]<br>Thickening [_]<br>Scarring [_]<br>Adhesion [_]<br>Normal peritoneum [_] | 0 [_]<br>1 [_]<br>2 [_]<br>3 [_] |
| 6 | Uterus [_]<br>Left Fallopian tube [_]<br>Right Fallopian tube [_]<br>Left ovary [_] | Tumour nodule [_]<br>Confluent nodules [_]<br>Plaque [_]<br>Thickening [_] | 0 [_]<br>1 [_]<br>2 [_]<br>3 [_] |

| | | | |
|---|---|---|---|
| | Right ovary [_]<br>Bladder [_]<br>Pouch of Douglas [_]<br>Rectosigmoid junction [_] | Scarring [_]<br>Adhesion [_]<br>Normal peritoneum [_] | |
| 7 | Right pelvic peritoneum [_]<br>Cecum [_]<br>Appendix [_]<br>Mesoappendix [_] | Tumour nodule [_]<br>Confluent nodules [_]<br>Plaque [_]<br>Thickening [_]<br>Scarring [_]<br>Adhesion [_]<br>Normal peritoneum [_] | 0 [_]<br>1 [_]<br>2 [_]<br>3 [_] |
| 8 | Right colon [_]<br>Right paracolic gutter [_]<br>Right mesocolon [_] | Tumour nodule [_]<br>Confluent nodules [_]<br>Plaque [_]<br>Thickening [_]<br>Scarring [_]<br>Adhesion [_]<br>Normal peritoneum [_] | 0 [_]<br>1 [_]<br>2 [_]<br>3 [_] |
| 9 | Proximal jejunum [_]<br>Proximal mesojejunum [_] | Tumour nodule [_]<br>Confluent nodules [_]<br>Plaque [_]<br>Thickening [_]<br>Scarring [_]<br>Adhesion [_]<br>Normal peritoneum [_] | 0 [_]<br>1 [_]<br>2 [_]<br>3 [_] |
| 10 | Distal jejunum [_]<br>Distal mesojejunum [_] | Tumour nodule [_]<br>Confluent nodules [_]<br>Plaque [_]<br>Thickening [_]<br>Scarring [_]<br>Adhesion [_]<br>Normal peritoneum [_] | 0 [_]<br>1 [_]<br>2 [_]<br>3 [_] |
| 11 | Proximal ileum [_]<br>Proximal mesoileum [_] | Tumour nodule [_]<br>Confluent nodules [_]<br>Plaque [_]<br>Thickening [_]<br>Scarring [_]<br>Adhesion [_]<br>Normal peritoneum [_] | 0 [_]<br>1 [_]<br>2 [_]<br>3 [_] |
| 12 | Distal ileum [_]<br>Distal ileum [_] | Tumour nodule [_]<br>Confluent nodules [_]<br>Plaque [_]<br>Thickening [_]<br>Scarring [_]<br>Adhesion [_]<br>Normal peritoneum [_] | 0 [_]<br>1 [_]<br>2 [_]<br>3 [_] |
| | Total | | |

Key points

- Each structure is considered only in one region. For e.g., a large omental cake may extend beyond region 0, but should be scored only in region 0
- When assigning the lesion score, the score of the structure bearing the largest tumor deposit should be considered and the morphology of that lesion scored

- The morphological features are defined as follows

1. **Tumor nodule** : Elevated mass with a round or irregular appearance
2. **Confluent nodules**: More than one nodule in a region with the intervening peritoneum showing abnormalities like smaller nodules or thickening. Alternatively, when the edges of discrete nodules become confluent.

3. **Plaque**: Elevated mass with a flat surface

4. **Thickening:** Thickened peritoneum without any of the above changes
5. **Scarring**- Lesion that has the appearance of scar tissue
6. **Adhesion**- Presence of adhesions between viscera or any viscera and the parietal peritoneum
7. **Normal peritoneum**- Absence of any of the changes described above
