## Supplement 4 for "Patterns of peritoneal dissemination and response to systemic chemotherapy in common and rare peritoneal tumors treated by cytoreductive surgery: Study protocol of a prospective, multi-center, observational study"

Pathological PCI
Select the structure with the largest tumor deposit in the region, select the type of lesion and the score (any one in each column)

| Region | Structure | Morphology (select any one) | Lesion score |
|---|---|---|---|
| 0 | Midline incision [_]<br>Anterior parietal peritoneum [_]<br>Greater omentum [_]<br>Omental cake [_]<br>Transverse colon [_]<br>Gastrocolic ligament [_]<br>Transverse mesocolon [_] | Tumour nodule [_]<br>Clustering of nodules [_]<br>Thickening [_]<br>Scarring [_]<br>Normal peritoneum [_] | 0 [_]<br>1 [_]<br>2 [_]<br>3 [_] |
| 1 | Right hepatic lobe surface [_]<br>Diaphragm [_]<br>Foramen of Winslow [_]<br>Gallbladder [_]<br>Hepatorenal recess [_] | Tumour nodule [_]<br>Clustering of nodules [_]<br>Thickening [_]<br>Scarring [_]<br>Normal peritoneum [_] | 0 [_]<br>1 [_]<br>2 [_]<br>3 [_] |
| 2 | Left hepatic lobe surface [_]<br>Lesser omentum [_]<br>Hepatic hilum [_]<br>Falciform ligament [_] | Tumour nodule [_]<br>Clustering of nodules [_]<br>Thickening [_]<br>Scarring [_]<br>Normal peritoneum [_] | 0 [_]<br>1 [_]<br>2 [_]<br>3 [_] |
| 3 | Surface of spleen [_]<br>Left diaphragmatic peritoneum [_]<br>Tail of pancreas and hilum of the spleen [_]<br>Anterior and posterior stomach surfaces [_] | Tumour nodule [_]<br>Clustering of nodules [_]<br>Thickening [_]<br>Scarring [_]<br>Normal peritoneum [_] | 0 [_]<br>1 [_]<br>2 [_]<br>3 [_] |
| 4 | Left colon [_]<br>Left paracolic gutter [_]<br>Left mesocolon [_] | Tumour nodule [_]<br>Clustering of nodules [_]<br>Thickening [_]<br>Scarring [_]<br>Normal peritoneum [_] | 0 [_]<br>1 [_]<br>2 [_]<br>3 [_] |
| 5 | Left pelvic peritoneum [_]<br>Secondary root of the mesosigmoid [_]<br>Sigmoid colon [_] | Tumour nodule [_]<br>Clustering of nodules [_]<br>Thickening [_]<br>Scarring [_]<br>Normal peritoneum [_] | 0 [_]<br>1 [_]<br>2 [_]<br>3 [_] |
| 6 | Uterus [_]<br>Left Fallopian tube [_]<br>Right Fallopian tube [_]<br>Left ovary [_]<br>Right ovary [_]<br>Bladder [_]<br>Pouch of Douglas [_]<br>Rectosigmoid junction [_] | Tumour nodule [_]<br>Clustering of nodules [_]<br>Thickening [_]<br>Scarring [_]<br>Normal peritoneum [_] | 0 [_]<br>1 [_]<br>2 [_]<br>3 [_] |
| 7 | Right pelvic peritoneum [_]<br>Cecum [_] | Tumour nodule [_]<br>Clustering of nodules [_] | 0 [_]<br>1 [_] |

| | | | |
|---|---|---|---|
| | Appendix [_]<br>Mesoappendix [_] | Thickening [_]<br>Scarring [_]<br>Normal peritoneum [_] | 2 [_]<br>3 [_] |
| 8 | Right colon [_]<br>Right paracolic gutter [_]<br>Right mesocolon [_] | Tumour nodule [_]<br>Clustering of nodules [_]<br>Thickening [_]<br>Scarring [_]<br>Normal peritoneum [_] | 0 [_]<br>1 [_]<br>2 [_]<br>3 [_] |
| 9 | Proximal jejunum [_]<br>Proximal mesojejunum [_] | Tumour nodule [_]<br>Clustering of nodules [_]<br>Thickening [_]<br>Scarring [_]<br>Normal peritoneum [_] | 0 [_]<br>1 [_]<br>2 [_]<br>3 [_] |
| 10 | Distal jejunum [_]<br>Distal mesojejunum [_] | Tumour nodule [_]<br>Clustering of nodules [_]<br>Thickening [_]<br>Scarring [_]<br>Normal peritoneum [_] | 0 [_]<br>1 [_]<br>2 [_]<br>3 [_] |
| 11 | Proximal ileum [_]<br>Proximal mesoileum [_] | Tumour nodule [_]<br>Clustering of nodules [_]<br>Thickening [_]<br>Scarring [_]<br>Normal peritoneum [_] | 0 [_]<br>1 [_]<br>2 [_]<br>3 [_] |
| 12 | Distal ileum [_]<br>Distal ileum [_] | Tumour nodule [_]<br>Clustering of nodules [_]<br>Thickening [_]<br>Scarring [_]<br>Normal peritoneum [_] | 0 [_]<br>1 [_]<br>2 [_]<br>3 [_] |
| | Total | | |

Key points

- Each structure is considered only in one region. For e.g., a large omental cake may extend beyond region 0, but should be scored only in region 0
- When assigning the lesion score, the score of the structure bearing the largest tumor deposit should be considered and the morphology of that lesion scored. The morphology is based on the findings of gross examination

- The morphological features are defined as follows

1. **Tumor nodule** : Elevated mass with a round or irregular appearance
2. **Clustering of nodules**: More than one nodule in a region with the intervening peritoneum showing abnormalities like smaller nodules or thickening. Alternatively, when the edges of discrete nodules become confluent.
3. **Scarring**- Lesion that has the appearance of scar tissue
4. **Adhesion**- Presence of adhesions between viscera or any viscera and the parietal peritoneum
5. **Normal peritoneum**- Absence of any of the changes described above
